# Dual-Model LLM Ensemble via Web Chat Interfaces Reaches Near-Perfect Sensitivity for Systematic-Review Screening: A Multi-Domain Validation with Equivalence to API Access

**DOI:** 10.1101/2025.11.03.25339455

**Authors:** Petter Fagerberg, Oscar Sallander, Kim Vikhe Patil, Anders Berg, Anastasia Nyman, Natalia Borg, Thomas Lindén

## Abstract

**Background:** Prior work showed that state-of-the-art (mid-2025) large language models (LLMs) prompted with varying batch sizes can perform well on systematic review (SR) abstract screening via public APIs within a single medical domain. Whether comparable performance holds when using no-code web interfaces (GUIs) and whether results generalize across medical domains remain unclear.

**Objective:** To evaluate the screening performance of a zero-shot, large-batch, two-model LLM ensemble (OpenAI GPT-5 Thinking; Google Gemini 2.5 Pro) operated via public chat GUIs across a diverse range of medical topics, and to compare its performance with an equivalent API-based workflow.

**Methods:** We conducted a retrospective evaluation using 736 titles and abstracts from 16 Cochrane reviews (330 included, 406 excluded), all published in May-June 2025. The primary outcome was the sensitivity of a pre-specified “OR” ensemble rule designed to maximize sensitivity, benchmarked against final full-text inclusion decisions (reference standard). Secondary outcomes were specificity, single-model performance, and duplicate-run reliability (Cohen’s κ). Because models saw only titles/abstracts while the reference standard reflected full-text decisions, specificity estimates are conservative for abstract-level screening.

**Results:** The GUI-based ensemble achieved 99.7% sensitivity (95% CI, 98.3%-100.0%) and 49.3% specificity (95% CI, 44.3%-54.2%). The API-based workflow yielded comparable performance, with 99.1% sensitivity (95% CI, 97.4%-99.8%) and 49.3% specificity (95% CI, 44.3%-54.2%). The difference in sensitivity was not statistically significant (McNemar p=0.625) and met equivalence within a ±2-percentage-point margin (TOST<0.05). Duplicate-run reliability was substantial to almost perfect (Cohen’s κ: 0.78-0.93). The two models showed complementary strengths: Gemini 2.5 Pro consistently achieved higher sensitivity (94.5%-98.2% across single runs), whereas GPT-5 Thinking yielded higher specificity (62.3%-67.0%).

**Conclusions:** A zero-code, browser-based workflow using a dual-LLM ensemble achieves near-perfect sensitivity for abstract screening across multiple medical domains, with performance equivalent to API-based methods. Ensemble approaches spanning two model families may mitigate model-specific blind spots. Prospective studies should quantify workload, cost, and operational feasibility in end-to-end systematic review pipelines.

## INTRODUCTION

Systematic reviews are the cornerstone of evidence-based medicine, forming the empirical basis for clinical guidelines, health policy, and critical treatment decisions [1]. However, their production is notoriously resource-intensive; a single review can require over a year of work [2] and cost more than €100,000 to complete [3]. A primary bottleneck in this process is title and abstract screening, which involves manually sifting through thousands of reports. The current gold standard, utilizing independent screening by two human reviewers [4], to maximize sensitivity and minimize bias [5] (especially important with less experienced reviewers [6, 7]), is so demanding that it is often infeasible to implement fully. Consequently, many teams resort to single-reviewer screening, which increases the risk of erroneously excluding relevant studies and introducing reviewer-specific bias. Furthermore, to manage this workload, research teams sometimes apply search filters that risk excluding relevant but poorly indexed studies. In this context, sensitivity is the dominant screening metric because missed eligible studies can bias effect estimates and guideline recommendations.

Large language models (LLMs) offer a promising solution to this screening bottleneck. While early investigations with older-generation LLMs demonstrated high sensitivity [8, 9], these approaches often required complex prompt engineering, API access, and custom code, creating a significant technical barrier for many review teams. More recent state-of-the-art models can achieve high performance with simpler, zero-shot prompting strategies, even when processing titles and abstracts in large batches [10]. Despite these advances, critical knowledge gaps persist. It remains unclear whether the high performance of these models, while prompted with large batches of titles and abstracts, is generalizable across diverse clinical domains. Furthermore, the run-to-run reliability of these models when accessed via public graphical user interfaces (GUIs), the modality most accessible to non-technical reviewers, has not been systematically evaluated.

To address these gaps, we evaluated a pragmatic, zero-code workflow for abstract screening. We deployed a two-model ensemble (OpenAI GPT-5 Thinking; Google Gemini 2.5 Pro) using simple, identical zero-shot prompts to screen the titles and abstracts from 16 diverse Cochrane reviews. The performance of this ensemble was validated against the final full-text inclusion/exclusion decisions made by the original human review teams.

Our primary objective was to evaluate the sensitivity of the LLM ensemble operated via public web chat GUIs and to compare its performance with an equivalent API-based workflow. Secondary outcomes included specificity, the performance of each constituent model, and the inter-run reliability of both the GUI-based approach and the API approach. We hypothesized that this accessible, browser-based workflow would achieve screening sensitivity comparable to that of more technically demanding API-based methods, thereby validating a potentially more practical tool for support during evidence synthesis.

## METHODS

### Study Design and Objectives

We conducted a retrospective, multi-domain validation study to evaluate the performance of large language models (LLMs) for systematic review abstract screening. The study compared two distinct operational modalities: 1) a no-code approach using web-based graphical user interfaces (GUIs), and 2) a programmatic approach using application programming interfaces (APIs). Our primary objective was to determine the screening sensitivity of a pre-specified two-model, two-run LLM ensemble. Secondary objectives were to evaluate the sensitivity and specificity of each individual model and to quantify the inter-run reliability (agreement between duplicate runs) for each model and modality. Analyses labeled pre-specified were defined before any model outputs were inspected; sensitivity analyses were exploratory.

### Data Source and Reference Standard

The validation dataset was derived from 16 diverse systematic reviews [11–26] published in the Cochrane Library (Issue 6, May-June 2025). The Cochrane reference standard was chosen due to its rigorous methodology, which includes independent screening by two human reviewers (the current gold standard), with a third human reviewer for final adjudication in cases of disagreements. For each Cochrane review, we extracted all titles and abstracts that the original authors had classified as either “included” or “excluded” following full-text assessment, through their provided EndNote libraries. Studies with indeterminate status (“ongoing” or “awaiting classification”) were omitted. We then programmatically retrieved the corresponding titles and abstracts in EndNote. Any citation for which an abstract could not be automatically retrieved was excluded from the final analysis dataset, as an abstract is required for the LLM screening task. The final dataset comprised 736 unique studies that had undergone full-text review, of which 348 were included and 388 were excluded by the Cochrane authors. The number of studies included per review ranged from 0 to 101, and the number of excluded studies ranged from 0 to 39.

### LLM Models and Access Modalities

We evaluated two state-of-the-art LLMs: OpenAI’s GPT-5 Thinking [27] and Google’s Gemini 2.5 Pro [28] due to their high performance in our previous study as well as other public benchmarks. For each model, we tested its performance via both its public web chat GUI and its public API. Google’s Gemini 2.5 Pro (API identifier gemini-2.5-pro) features a knowledge cutoff of January 2025 and an API input token limit of 1,048,576. In comparison, OpenAI’s GPT-5 Thinking (gpt-5-2025-08-07) has an October 2024 knowledge cutoff and a 400,000-input token limit. It is important to specify that these token limits are for the API versions; the corresponding limits for the GUI versions are unknown. We configured API settings to align with the default settings (Gemini: temperature = 1.0; OpenAI: reasoning effort = medium). For each run, we initiated a fresh chat/session with no prior context.

### Prompting and Screening Workflow

We employed a zero-shot system prompt that instructed the LLM to function as an expert systematic reviewer screening titles and abstracts against eligibility criteria (see our previous publication for more details [10]). We extracted PICOS (Population, Intervention, Comparator, Outcomes, Study design) eligibility criteria from each Cochrane review’s methods section. The prompt directed the model to classify each citation as include, exclude, or full-text review (in cases of uncertainty), provide a single-sentence justification, and format the output as a machine-readable markdown table. Screening was performed by providing large batches of titles and abstracts to the models (Table 1). For reviews with ≤100 records, we screened in one batch; for those with >100, we split them into two batches to remain under model token limits applicable in the GUI versions of each model. To handle occasional technical failures (i.e., non-completed output), the batch was run in a fresh session to avoid context contamination.

**Table 1.**
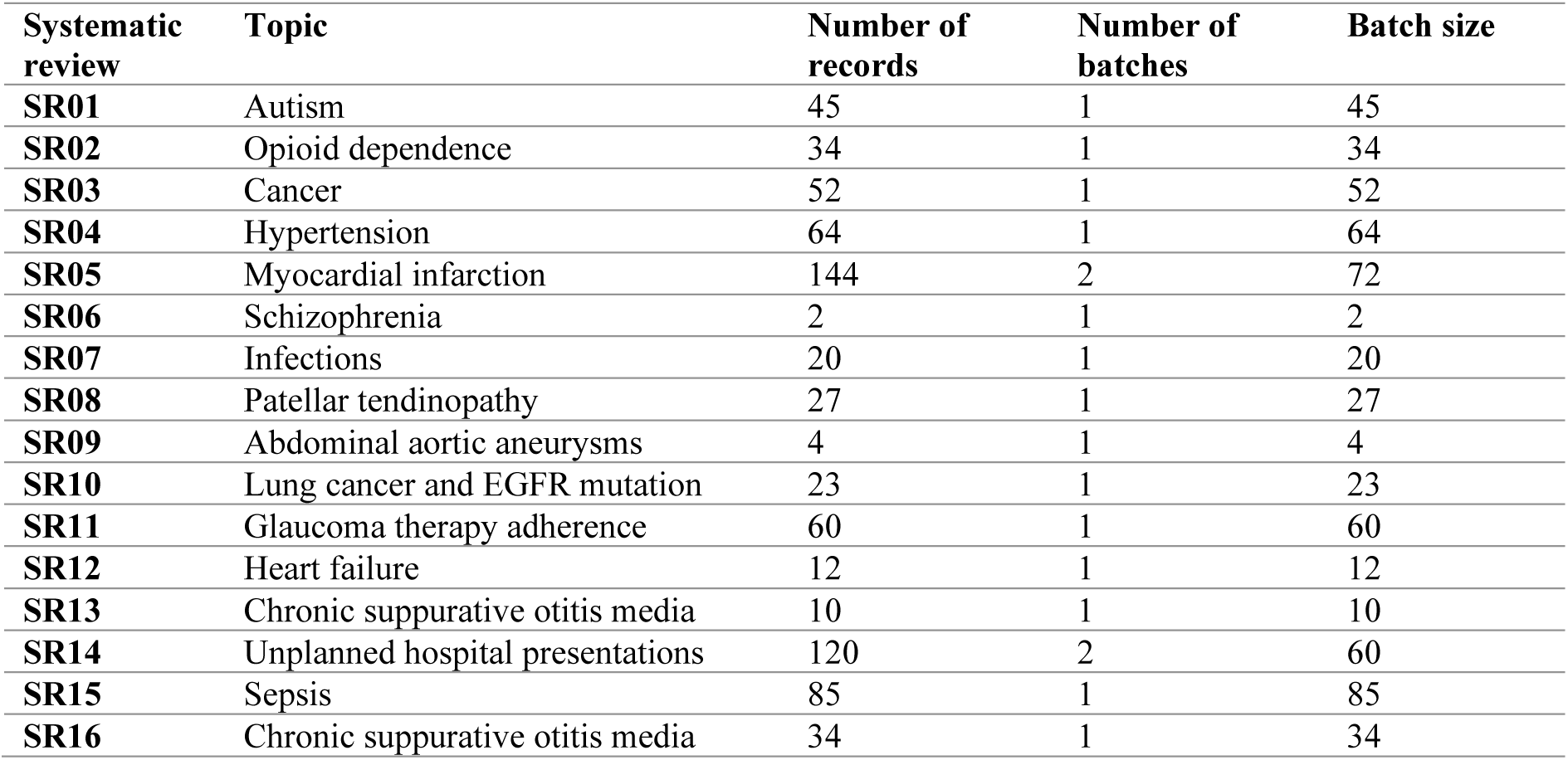
The prompt level batch size (number of titles and abstracts per prompt) that was used for each systematic review [11–26].

| Systematic review | Topic | Number of records | Number of batches | Batch size |
| --- | --- | --- | --- | --- |
| SR01 | Autism | 45 | 1 | 45 |
| SR02 | Opioid dependence | 34 | 1 | 34 |
| SR03 | Cancer | 52 | 1 | 52 |
| SR04 | Hypertension | 64 | 1 | 64 |
| SR05 | Myocardial infarction | 144 | 2 | 72 |
| SR06 | Schizophrenia | 2 | 1 | 2 |
| SR07 | Infections | 20 | 1 | 20 |
| SR08 | Patellar tendinopathy | 27 | 1 | 27 |
| SR09 | Abdominal aortic aneurysms | 4 | 1 | 4 |
| SR10 | Lung cancer and EGFR mutation | 23 | 1 | 23 |
| SR11 | Glaucoma therapy adherence | 60 | 1 | 60 |
| SR12 | Heart failure | 12 | 1 | 12 |
| SR13 | Chronic suppurative otitis media | 10 | 1 | 10 |
| SR14 | Unplanned hospital presentations | 120 | 2 | 60 |
| SR15 | Sepsis | 85 | 1 | 85 |
| SR16 | Chronic suppurative otitis media | 34 | 1 | 34 |

### Experimental Protocol and Ensemble Definition

The entire dataset of 736 titles and abstracts was screened by each model four times: two independent, duplicate runs via the GUI and two duplicate runs via the API. This 2 models × 2 runs × 2 modalities design yielded paired duplicate runs for each model within each access condition. For the primary analysis, we defined a highly inclusive ensemble decision rule. Within each modality (GUI and API), the ensemble consisted of all four corresponding runs (two from GPT-5, two from Gemini 2.5 Pro). A citation was classified as positive if any of the four runs labeled it “Include” or “Full-text”. This “OR” logic was chosen to maximize overall sensitivity.

### Outcomes and Statistical Analysis

The primary outcome was the sensitivity of the ensemble’s binary classification (include vs. exclude) relative to the reference standard. We mapped “include” and “full-text” to positive and “exclude” to negative for all primary and reliability analyses. Secondary outcomes included sensitivity and specificity for each individual model run. We computed exact (Clopper-Pearson) 95% CIs for proportions, used McNemar tests [29] for paired comparisons, and conducted a TOST equivalence test on sensitivity with a ±2-point margin [30]. Inter-run reliability for duplicate runs was quantified using Cohen’s κ coefficient [31, 32], with 95% CIs generated via bootstrapping (2000 replicates). All analyses were conducted in R (version 4.5.1 [33]).

### Targeted Adjudication of Discrepancies

To explore the nature of critical disagreements between the LLMs and the reference standard, we implemented a pre-specified targeted adjudication protocol. This analysis was designed to understand potential LLM failure modes as well as correcting potential reference standard decisions that were discordant with the eligibility criteria. A citation was flagged for adjudication if: 1) both duplicate runs of the two LLMs disagreed with the reference label, or 2) both duplicate runs of both models classified a record as “exclude”, while Cochrane also had labeled it as “exclude”. The second condition was used because every citation in our dataset was deemed relevant enough for full-text review by the original authors, making any consistent LLM-based exclusion a high-priority event to investigate.

Two adjudicators, trained in the systematic review process, were provided with the flagged titles/abstracts and the review’s eligibility criteria. Disagreements were resolved by consensus. This adjudication was performed to characterize errors made by the LLMs as well as to better align the reference standard with the actual criteria of each original Cochrane review. The primary outcome results are presented with this improved reference standard and results vs. the original Cochrane EndNote libraries are provided with supplementary material.

18 records (2.3%) met adjudication criterion 1, and 23 (3.1%) met criterion 2. All 18 criterion 1 records were judged to be probable misclassifications in the original EndNote libraries when evaluated against the prespecified review criteria. Of the 23 criterion 2 records, which were initially sent for full-text assessment by Cochrane authors but later excluded, only one had an abstract sufficiently ambiguous that, based on title and abstract information alone, it should have been advanced to full-text review. On full-text assessment, this study was correctly excluded. Nonetheless, we cannot rule out that AI models may have been trained on the full-text content of this paper, because the article was openly accessible rather than behind a paywall. Overall, the discrepancies appear to stem primarily from library misclassification rather than screening errors. The final gold standard dataset after the adjudication process contained 330 records labeled as “include” and 406 “exclude”. The final gold standard was used for all comparisons.

## RESULTS

### Primary Outcome: Ensemble Performance

The pre-specified two-model, two-run ensemble demonstrated high sensitivity in both operational modalities. In the code-free GUI condition, the ensemble identified 329 of 330 included studies, achieving 99.7% sensitivity (95% CI, 98.3%-100.0%) and 49.3% specificity (95% CI, 44.3%-54.2%) (Figure 2). Performance in the programmatic API condition was nearly identical, yielding 99.1% sensitivity (95% CI, 97.4%-99.8%) by identifying 327 of 330 included studies, with the same specificity of 49.3% (95% CI, 44.3%-54.2%). McNemar testing indicated no significant difference (p = 0.625). The TOST procedure supported equivalence within ±2 percentage points (paired; p lower = 1.1×10⁻⁵, p upper = 0.011). As expected from the inclusive “OR” ensemble rule, false negatives were rare: the GUI ensemble missed 1/330 (0.3%) and the API ensemble 3/330 (0.9%).

**Figure 1.**
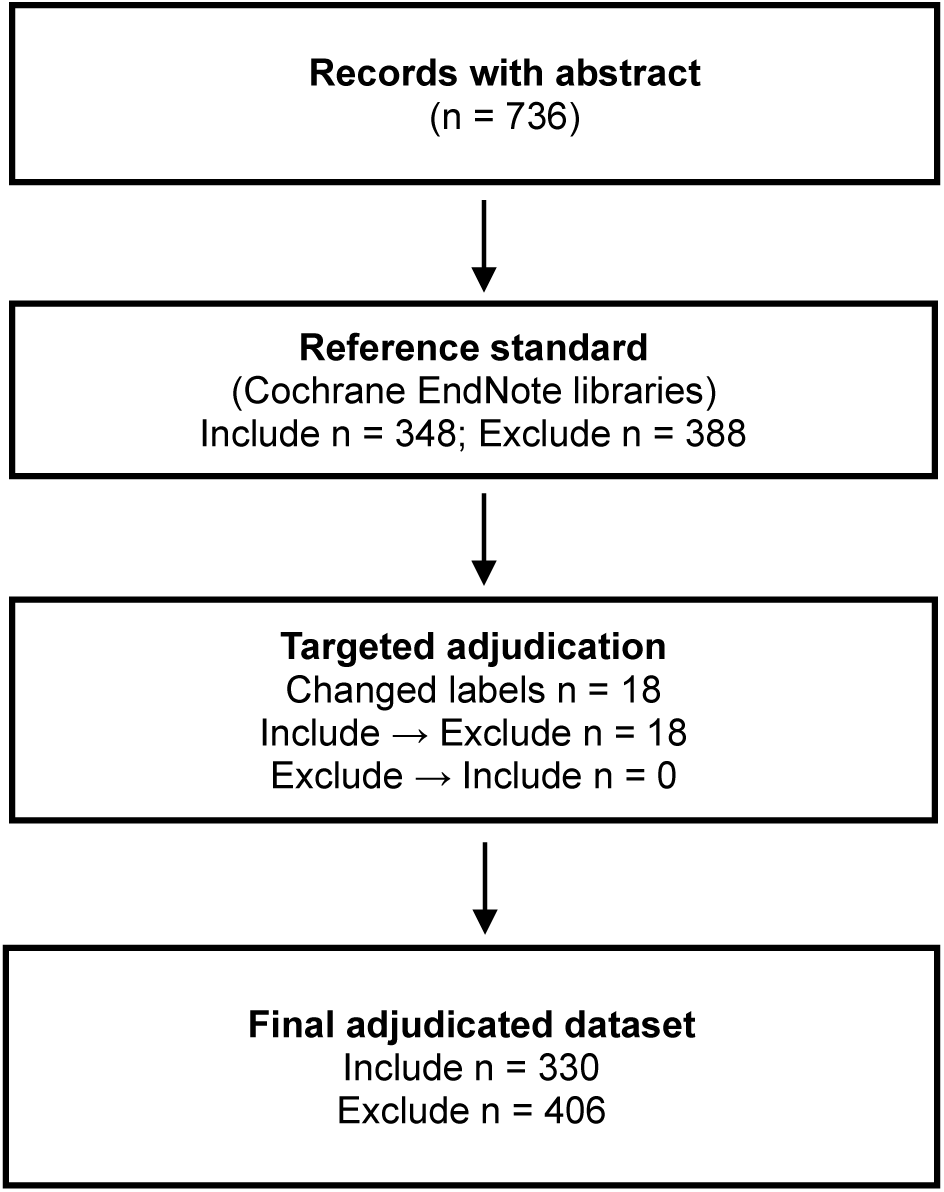
Study flow diagram showing the derivation of reference standard. Records progressed from initial Cochrane EndNote library classifications through targeted adjudication to create the final gold standard dataset. Numbers indicate studies at each stage with reason for classification changes.

**Figure 2.**
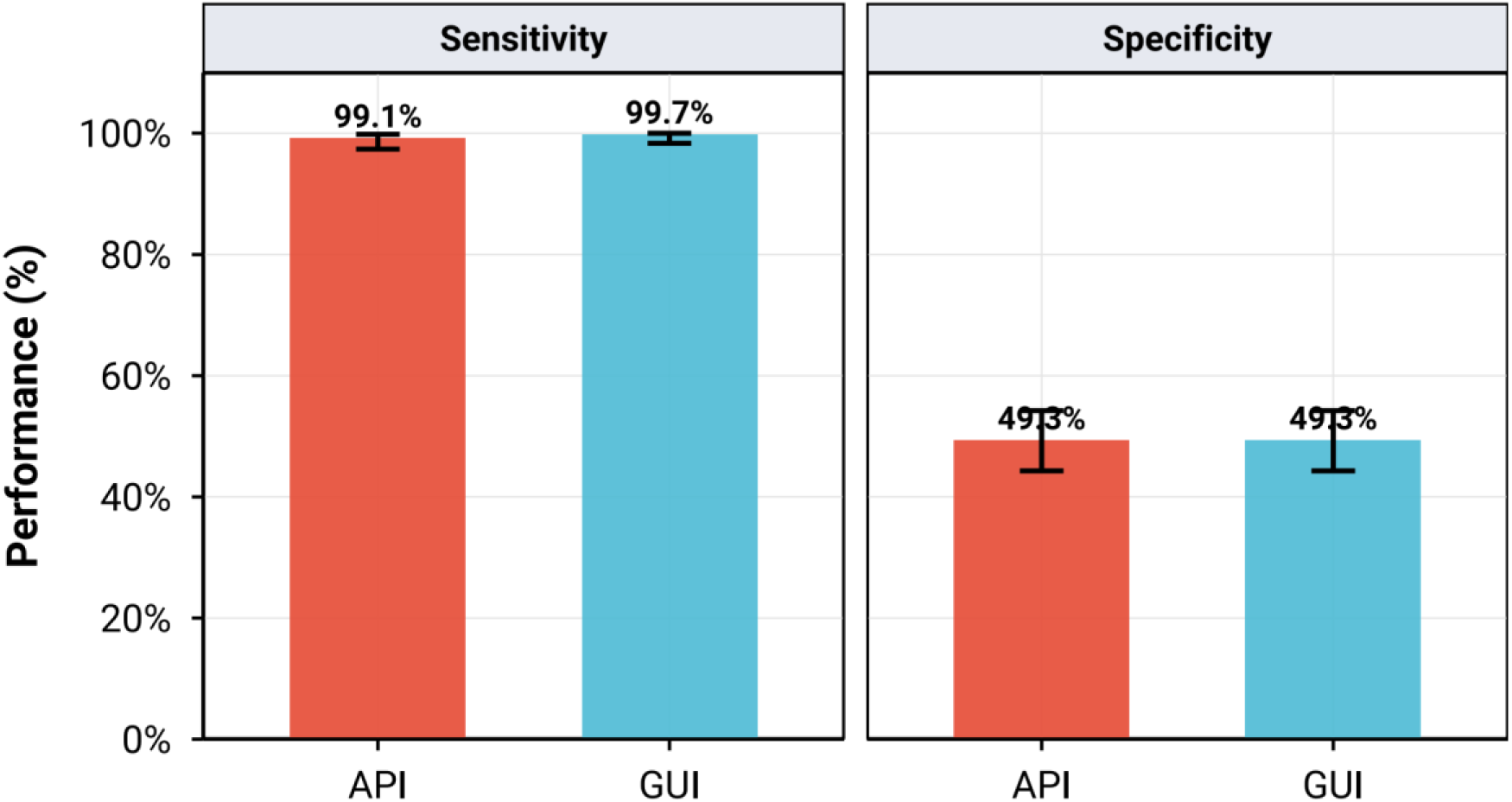
Performance of the four-run ensemble. Sensitivity and specificity of the pre-specified two-model, two-run ensemble using an “OR” decision rule across the full dataset (N=736 titles and abstracts). Error bars represent 95% Clopper-Pearson confidence intervals.

### Single-Run Ensemble Performance

A pre-specified sensitivity analysis of a simpler ensemble (one run per model) showed that specificity increases with only a small decrease in sensitivity. Using run 1 for each model, the GUI single-run ensemble achieved 99.1% sensitivity (327/330) with 56.2% specificity (95% CI, 51.2%-61.0%), while the API single-run ensemble achieved 98.5% sensitivity (325/330) with 53.9% specificity (95% CI, 49.0%-58.9%).

### Single-Model Performance

Analysis of individual runs revealed complementary profiles underlying the ensemble’s performance (Figure 3 and Figure 4). Gemini 2.5 Pro consistently achieved higher sensitivity across both GUI and API modalities (94.5%-98.2% across runs), whereas GPT-5 Thinking consistently yielded higher specificity (65.0%-67.0%). Detailed per-run metrics are provided in the supplementary material.

**Figure 3.**
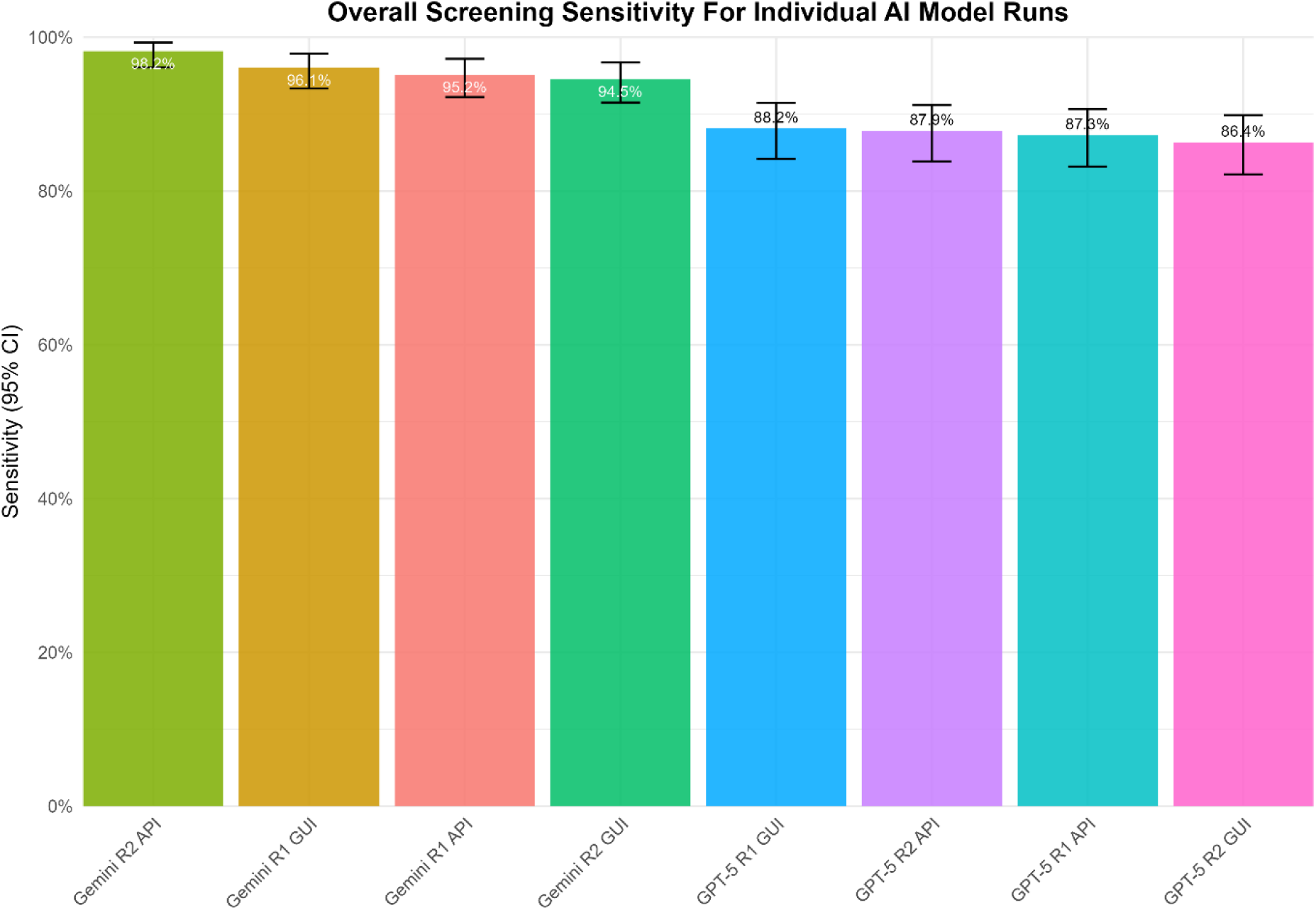
Individual model run sensitivity. Screening sensitivity for each of the eight individual runs across the full dataset (N=736). Error bars represent 95% Clopper-Pearson confidence intervals.

**Figure 4.**
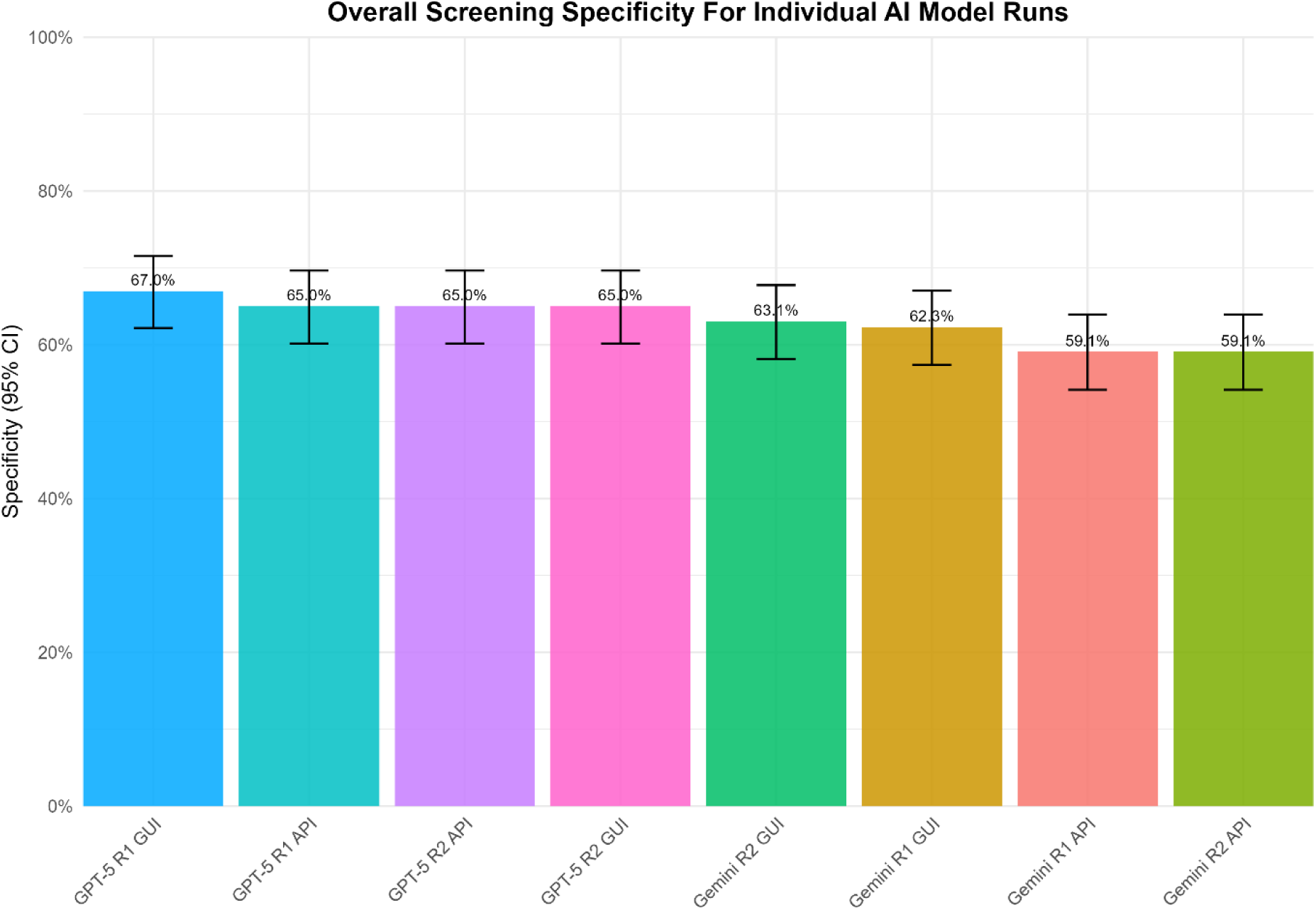
Individual model run specificity. Screening specificity for each of the eight individual runs across the full dataset (N=736). Error bars represent 95% Clopper-Pearson confidence intervals.

### Duplicate-Run Reliability

Both models demonstrated a high degree of stochastic consistency between duplicate runs. Inter-run reliability was substantial to almost perfect across all model-modality pairings, with Cohen’s kappa (κ) values ranging from 0.78 to 0.93 (Figure 5). These findings indicate that for a given prompt, the models produce highly reliable classifications from one run to the next.

**Figure 5.**
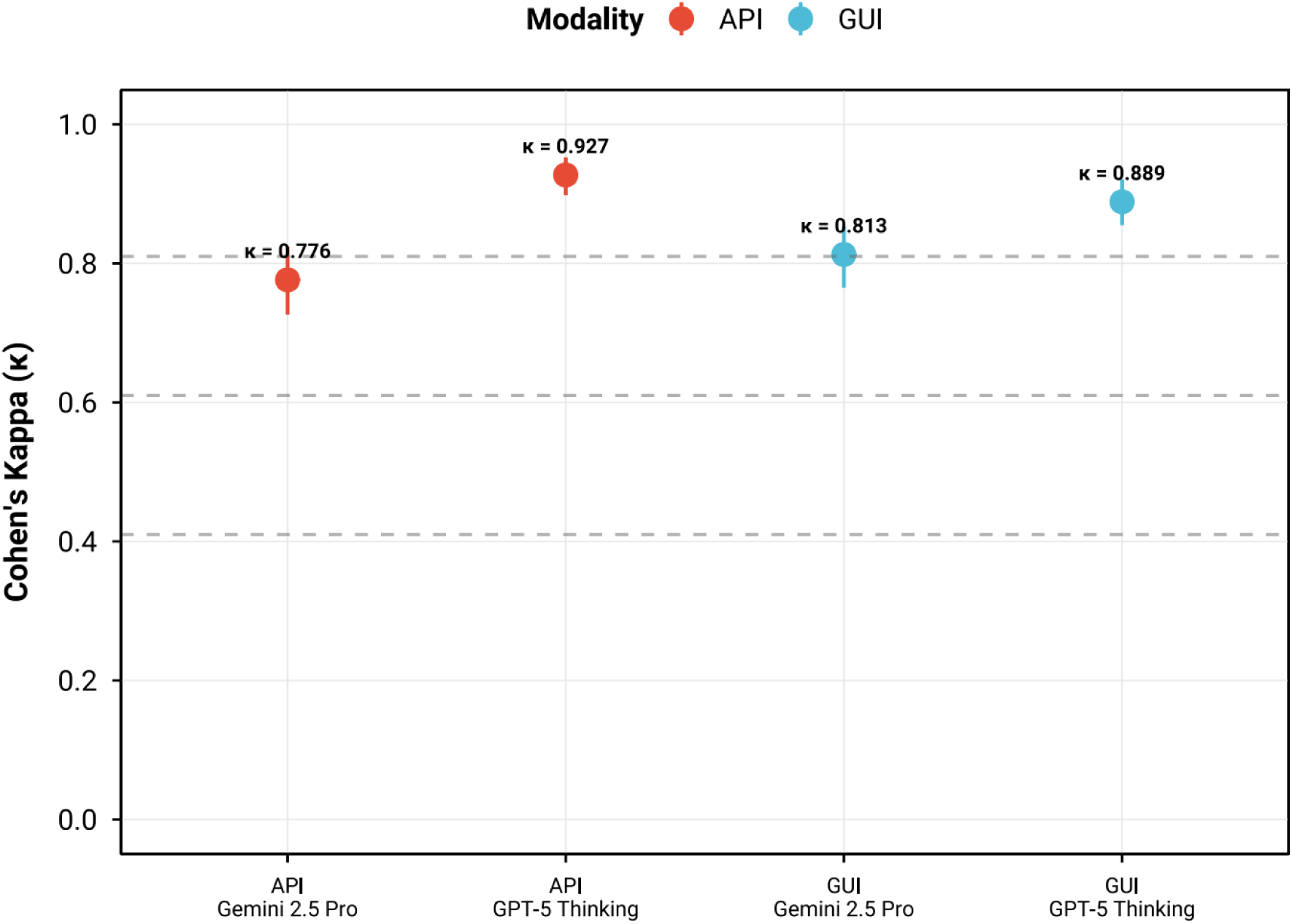
Duplicate-Run Reliability. Inter-run reliability (Cohen’s κ) for duplicate screening runs (N=736). The dashed line at κ = 0.81 indicates the threshold for “almost perfect agreement”, at κ = 0.61 “substantial agreement”, and at κ = 0.41 ”moderate agreement” [31]. Error bars represent 95% bootstrap confidence intervals.

### Performance Across Clinical Domains

While overall performance was robust for the ensemble, we observed some variability across the 16 different Cochrane reviews within individual runs of each model, suggesting certain clinical topics were more challenging for the models. For instance, GPT-5 Thinking single-run sensitivity was lowest (43%, 3/7 true positives) in systematic review 7 (Vaccines for preventing infections in adults with haematological malignancies [17]), suggesting model specific blind spots and the value of ensembling. Reviews with very few positives displayed wide uncertainty (i.e., some SRs had ≤3 included studies), as reflected in broad confidence intervals. Per-review performance data are available in supplementary material.

## DISCUSSION

Our study makes two key contributions. First, we demonstrate the cross-domain generalizability of a zero-shot large batch title and abstract screening approach by testing it on a diverse corpus of 16 Cochrane reviews. The two-model LLM ensemble achieved near-perfect sensitivity (99.1%-99.7%) on this task. Second, we provide the first direct comparison of public web GUIs (99.7%) versus programmatic APIs (99.1%), confirming that comparable, high-sensitivity results can be achieved without programming expertise.

As designed, our inclusive ensemble rule prioritized sensitivity at the expense of specificity, a trade-off confirmed by analysis of the constituent models; Gemini 2.5 Pro consistently provided higher sensitivity, while GPT-5 Thinking yielded higher specificity. These complementary profiles justify a cross-family ensemble. Importantly, we observed that individual models exhibited unique failure modes on certain reviews, underscoring the importance of the ensemble approach to mitigate model-specific biases and reduce the risk of idiosyncratic sensitivity failures. For example, a post-hoc examination of the most challenging review for the LLMs (vaccines in haematological populations [17]) provided a clear example of a model-specific failure mode. GPT-5 Thinking systematically excluded studies involving the general population, incorrectly assuming the target subgroup (patients with haematological malignancies) was absent. This represents an over-confident reasoning error. In contrast, Gemini 2.5 Pro demonstrated a more cautious approach, correctly identifying the ambiguity and flagging these same records for full-text assessment to confirm the presence of the subgroup. Prompt engineering techniques, such as using few-shot examples of erroneous exclusions, could be a viable method to steer GPT-5 towards a more cautious screening approach. While we cannot fully generalize this specific behavior from our limited samples, a cross-family ensemble provides an important safeguard. In this case, one model’s caution compensates for another’s confident error, directly mirroring the rationale for using dual human reviewers to mitigate individual human bias.

Because performance was benchmarked against final full-text inclusion/exclusion decisions, this dataset represents a particularly challenging benchmark for abstract screening. Thus, the obtained specificities represent conservative lower bounds for real-world abstract-level screening. Furthermore, both models demonstrated substantial to almost perfect inter-run reliability (Cohen’s κ: 0.78-0.93), indicating a high degree of stochastic stability.

The efficiency of LLM-based screening has important implications for the initial search strategy. To manage workload, review teams may apply methodological search filters that, while greatly reducing the volume of scientific papers, carry the risk of systematically excluding relevant studies that are poorly indexed. Because the LLM workflow is not constrained by human time in the same way, it can reduce the need for more aggressive filtering. Researchers can instead use broader, more sensitive search strategies and allow the LLM ensemble to perform the initial comprehensive triage in the broader list of studies. This capability can serve as an important protection layer, enhancing the completeness of a review by mitigating the risk of inadvertently omitting studies at the search stage. Additionally, exclusions made by the ensemble at the abstract level, save the time and cost associated with retrieving and assessing full-text articles that would have been excluded anyway.

For systematic reviews, failing to identify an eligible study (a false negative) is typically the most critical error [4]. Previous studies utilizing a single human screener estimated overall human performance at 87-92% sensitivity (reported range 42-100% for individual reviewers) as a comparison [6, 7], highlighting the risk of missing relevant studies with such approach. Our findings therefore validate a practical, zero-code workflow that rigorously minimizes this risk. This work opens the door for several models of human-AI collaboration. First, in the gold-standard dual-reviewer workflow, the LLM ensemble could act as a decision-support tool, providing a third opinion to resolve initial human disagreements and potentially reducing the burden on a senior adjudicator. Second, for resource-constrained teams unable to perform dual review, the LLM ensemble could serve as a robust second screener alongside a single human reviewer. This approach could significantly reduce the risk of error associated with single-reviewer screening while requiring only half the human effort of the traditional gold standard. Lastly, a semi-automated workflow could involve the LLM ensemble performing the initial screen of all studies. A human reviewer would then assess all records flagged for inclusion or full-text review, plus a small, random sample of the excluded studies to safeguard against systematic model bias (e.g., 5-10% or balanced with the number of inclusions). This approach enables monitoring of emerging failure modes and concept drift. Our finding that individual models had unique blind spots underscores the continued importance of a human-in-the-loop to ensure accountability.

While LLMs can potentially assist at multiple stages of a systematic review [34], i.e., from search strategy formulation to data extraction and risk of bias assessment, title and abstract screening represents a more accessible entry point. This is because titles and abstracts are publicly available bibliographic data. In contrast, subsequent steps like data extraction or quality appraisal require processing of full-text content. Therefore, abstract screening might be a more pragmatic initial proof-of-concept for LLM adoption within the systematic review process.

A central implication of our work is the validation of public web GUIs as a technically viable access point for high-sensitivity abstract screening. This finding greatly reduces the barrier to adoption for research teams that lack the specialized programming expertise or infrastructure required for API-based methods, thereby helping to democratize the use of AI in evidence synthesis. However, this accessibility is coupled with significant governance challenges. Public-facing models are subject to unannounced updates (“model drift”), which may compromise scientific reproducibility over time. The ensemble approach itself offers a degree of resilience against such challenges; by diversifying across two model families from different providers, the risk of a single silent update compromising an entire workflow is mitigated. To address these challenges further, a framework for responsible implementation might include: i) using fixed model versions or snapshots with documented run dates; ii) ensuring session isolation and disabling external tool access (e.g., browsing); iii) securing institutional data processing agreements; iv) maintaining comprehensive audit logs of all prompts and outputs; and v) conducting periodic performance checks against a frozen benchmark dataset to detect model drift. Furthermore, the human-computer interaction aspect is also crucial when evaluating the use of a GUI for this process. The accuracy of prompt engineering and data handling directly governs the validity of the LLM’s output. For example, simple user errors during manual data transfer, such as faulty copy-pasting, can lead to highly problematic or entirely incorrect outcomes. Therefore, implementing rigorous training protocols and safeguards is essential to support novices screening systematic reviews with this new methodology. To minimize such user-induced errors, the optimal long-term approach may be a fully automated, API-based pipeline featuring a purpose-built, simplified interface, which offers greater reliability than open-ended public LLM GUIs. A real-world implementation might therefore still need to trend towards enterprise-grade, privacy-preserving platforms (e.g., Google Vertex AI [35], OpenAI API platform [36]) that offer better versioning, security, and regulatory compliance.

Another alternative would be to extend this validation to high-performing open-source LLMs, which offer the potential for local deployment. For government agencies, healthcare systems, and other organizations, a locally hosted ensemble would mitigate many of the governance challenges inherent to commercial cloud services. It would ensure stable, consistent performance by allowing teams to lock in a specific model version, thus avoiding performance drift. Furthermore, it provides a more robust approach for navigating the complex and evolving legal landscape surrounding data privacy and sovereignty. Beyond governance, local deployment could unlock the capability to fine-tune models using expert human adjudication decisions as training data, creating a feedback loop to correct systematic model errors and progressively enhance screening accuracy over time. Furthermore, local deployment would enable the use of LLMs for downstream systematic review tasks, such as data extraction and quality appraisal, which are currently unfeasible with third-party APIs due to further restrictions on processing full-text articles.

Our findings should be interpreted in the context of several limitations. First, our reference standard was the final full-text inclusion decision, whereas the LLMs screened only titles and abstracts. This design choice robustly tests sensitivity but yields a conservatively low estimate of specificity. Many abstracts correctly excluded by the LLMs would also have been excluded at the abstract screening stage by human reviewers, but our methodology counts these as false positives against the full-text standard. Relatedly, our dataset comprised only studies that had already survived an initial human screening to be considered for full-text review, representing a set of uniquely challenging records. Real-world specificity on a completely unscreened search result set would therefore likely be higher, as demonstrated in our prior study [10]. Second, methodological constraints prevented perfect parity between the GUI and API environments. Stochastic settings like temperature could not be fully harmonized, and we could not rule out minor, unannounced vendor-side updates during the study period, although the high inter-run reliability suggests this had minimal impact. Third, our analysis did not measure operational factors such as cost, processing time (throughput), or user acceptability, which are critical for real-world implementation and should be assessed in prospective studies. Fourth, a potential for data contamination exists if the Cochrane reviews used for this validation were part of the models’ pre-training corpora, although Cochrane reviews published in May 2025 after AI model training mitigated this risk. Finally, while robust across 16 diverse reviews, our analysis may not capture all rare edge cases.

Beyond augmenting the screening process itself, our findings suggest a novel application for LLMs in the foundational stages of systematic review protocol development. Traditionally, formulating PICOS criteria is an arduous process, and flaws may only become apparent after substantial human screening effort has been invested. LLMs could facilitate a paradigm of rapid, iterative protocol testing; a review team could draft PICOS criteria, conduct a preliminary search, and immediately deploy an LLM ensemble to see which studies are included. This provides immediate, concrete feedback on the criteria’s real-world performance, allowing for rapid refinement before committing to full-scale human review. Also, author teams could test database searches with and without search filters to see if there is a high risk of losing important studies when applying the filters. By lowering the barrier to iterative testing and sensitive search approaches, the quality of systematic reviews could be improved.

This work also establishes the foundation for prospective, end-to-end integration studies. Future research should quantify the real-world impact of these workflows on reviewer workload, project timelines, and cost-effectiveness. Key technical questions remain, including the optimization of ensemble strategies and the development of risk-calibrated workflows that route only the most uncertain cases to human experts. Critically, the research community should try to agree on robust governance frameworks and reporting standards to guide the ethical and reproducible use of LLMs in evidence synthesis.

## CONCLUSION

A zero-code, browser-based workflow using a dual-LLM ensemble can achieve near-perfect sensitivity for systematic review abstract screening across diverse medical topics, with performance statistically non-inferior to an API-based workflow. These findings validate an accessible approach that can augment review teams, support novel applications like iterative protocol refinement, and ultimately accelerate the pace of evidence synthesis. The use of a cross-family ensemble was shown to mitigate the risks of model-specific biases and maximize sensitivity. The successful integration of these tools will now depend on quantifying their operational benefits and establishing robust governance models, including the exploration of locally deployed open-source alternatives, that ensure security, privacy, and scientific reproducibility.

## Acknowledgements

We thank the authors of the original Cochrane Reviews for their rigorous work and for making their EndNote libraries available.

## Ethics approval and consent to participate

Not applicable.

## Author contributions

PF and OS had full access to all the data in the study and took responsibility for the integrity of the data and the data analysis. Concept and design: PF, OS and KVP. Acquisition, analysis, or interpretation of data: PF and OS. Drafting of the manuscript: PF, KVP and OS. Critical review of the manuscript for important intellectual content: All authors. Statistical analysis: PF and OS. Administrative, technical, or material support: AB, AN, TL, NB. Supervision: AB, AN, TL, NB. All authors read and approved the definitive version of the manuscript.

## Data availability statement

The datasets used and analysed in this study are available upon reasonable request.

## SUPPLEMENTARY MATERIAL

**Complete List of Cochrane Reviews Included as Reference Standard Material [11–26]**

SR1. Atypical antipsychotics for autism spectrum disorder: a network meta-analysis

SR2. Sustained-release naltrexone for opioid dependence

SR3. Yoga for fatigue in people with cancer

SR4. Dietary Approaches to Stop Hypertension (DASH) for the primary and secondary prevention of cardiovascular diseases

SR5. Stem cell treatment for acute myocardial infarction

SR6. Post-incident debriefing for people with schizophrenia after coercive measures

SR7. Vaccines for preventing infections in adults with haematological malignancies SR8. Exercise for patellar tendinopathy

SR9. Fenestrated endovascular repair for abdominal aortic aneurysms

SR10. Adjuvant epidermal growth factor receptor (EGFR) tyrosine kinase inhibitors (TKIs) for the treatment of people with resected stage I to III non-small-cell lung cancer and EGFR mutation

SR11. Multifaceted behavioral interventions to improve topical glaucoma therapy adherence in adults

SR12. Antiplatelet versus anticoagulation treatment for people with heart failure in sinus rhythm

SR13. Aural toilet (ear cleaning) for chronic suppurative otitis media

SR14. Community care navigation intervention for people who are at risk of unplanned hospital presentations

SR15. Corticosteroids for treating sepsis in children and adults

SR16. Antibiotics versus topical antiseptics for chronic suppurative otitis media

## LLM Performance vs. Corrected Cochrane EndNote library

**Table S1.**
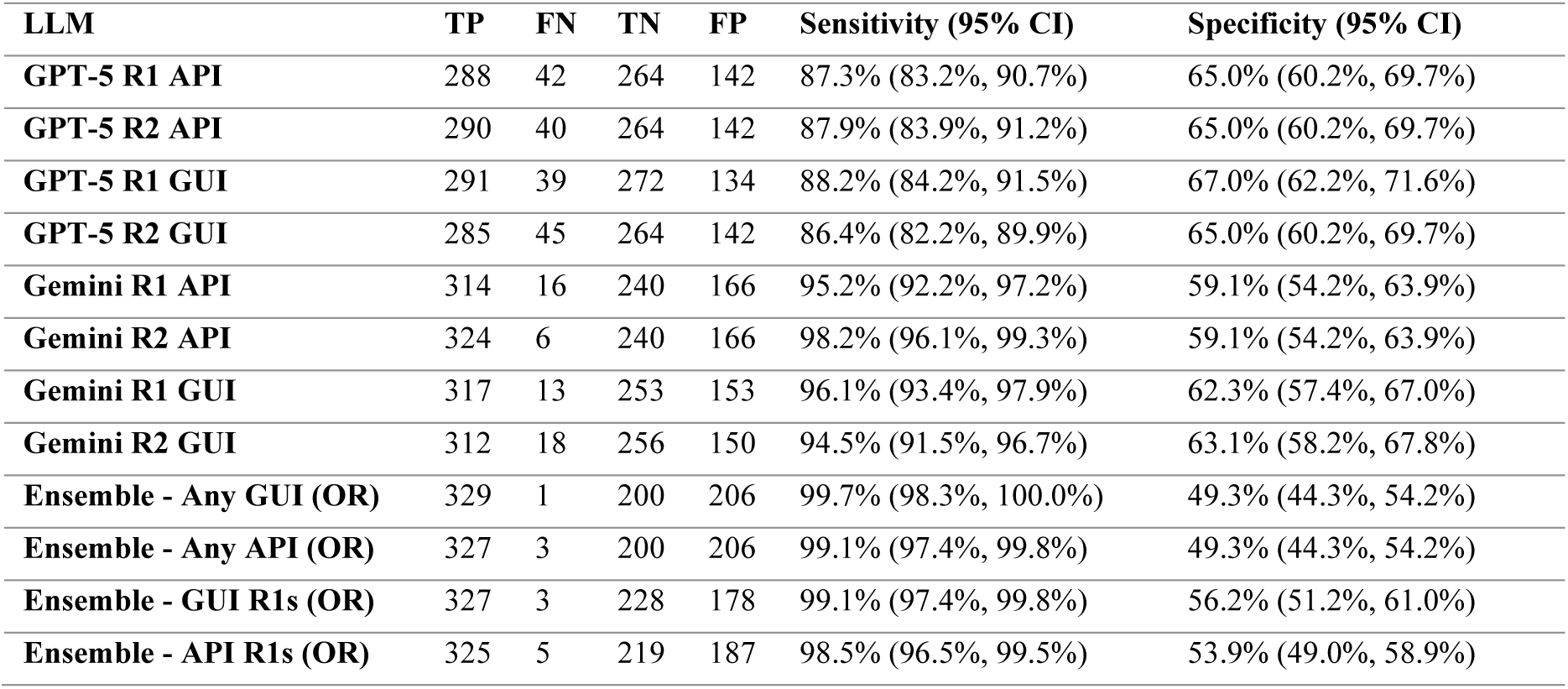
Overall Performance of All Model Variants. Summary of performance metrics, including true positives (TP), false negatives (FN), true negatives (TN), false positives (FP), sensitivity, and specificity. Confidence intervals (CI) are 95% Clopper-Pearson intervals.

| LLM | TP | FN | TN | FP | Sensitivity (95% CI) | Specificity (95% CI) |
| --- | --- | --- | --- | --- | --- | --- |
| GPT-5 R1 API | 288 | 42 | 264 | 142 | 87.3% (83.2%, 90.7%) | 65.0% (60.2%, 69.7%) |
| GPT-5 R2 API | 290 | 40 | 264 | 142 | 87.9% (83.9%, 91.2%) | 65.0% (60.2%, 69.7%) |
| GPT-5 R1 GUI | 291 | 39 | 272 | 134 | 88.2% (84.2%, 91.5%) | 67.0% (62.2%, 71.6%) |
| GPT-5 R2 GUI | 285 | 45 | 264 | 142 | 86.4% (82.2%, 89.9%) | 65.0% (60.2%, 69.7%) |
| Gemini R1 API | 314 | 16 | 240 | 166 | 95.2% (92.2%, 97.2%) | 59.1% (54.2%, 63.9%) |
| Gemini R2 API | 324 | 6 | 240 | 166 | 98.2% (96.1%, 99.3%) | 59.1% (54.2%, 63.9%) |
| Gemini R1 GUI | 317 | 13 | 253 | 153 | 96.1% (93.4%, 97.9%) | 62.3% (57.4%, 67.0%) |
| Gemini R2 GUI | 312 | 18 | 256 | 150 | 94.5% (91.5%, 96.7%) | 63.1% (58.2%, 67.8%) |
| Ensemble - Any GUI (OR) | 329 | 1 | 200 | 206 | 99.7% (98.3%, 100.0%) | 49.3% (44.3%, 54.2%) |
| Ensemble - Any API (OR) | 327 | 3 | 200 | 206 | 99.1% (97.4%, 99.8%) | 49.3% (44.3%, 54.2%) |
| Ensemble - GUI R1s (OR) | 327 | 3 | 228 | 178 | 99.1% (97.4%, 99.8%) | 56.2% (51.2%, 61.0%) |
| Ensemble - API R1s (OR) | 325 | 5 | 219 | 187 | 98.5% (96.5%, 99.5%) | 53.9% (49.0%, 58.9%) |

**Supplementary Figure S1-S12:**
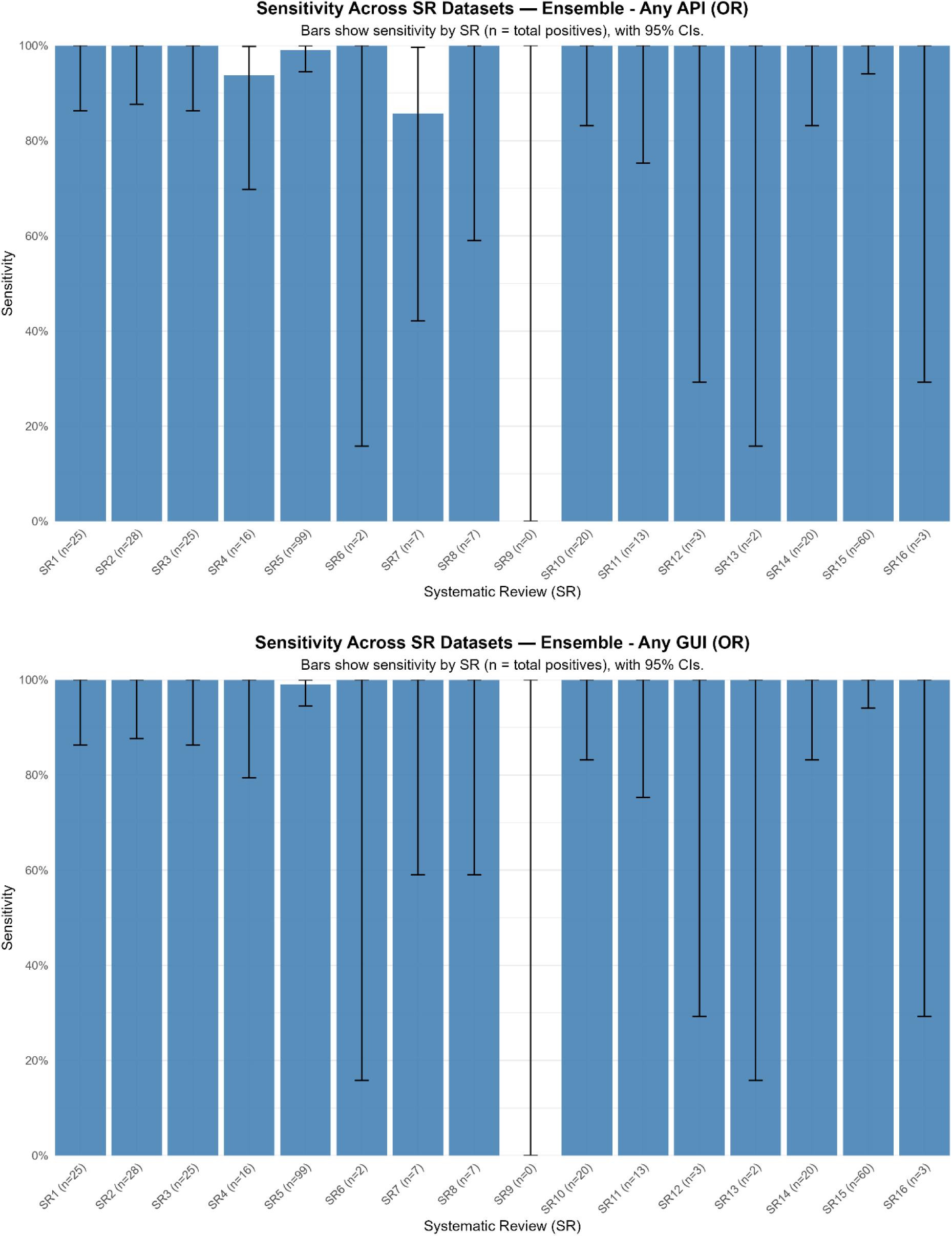
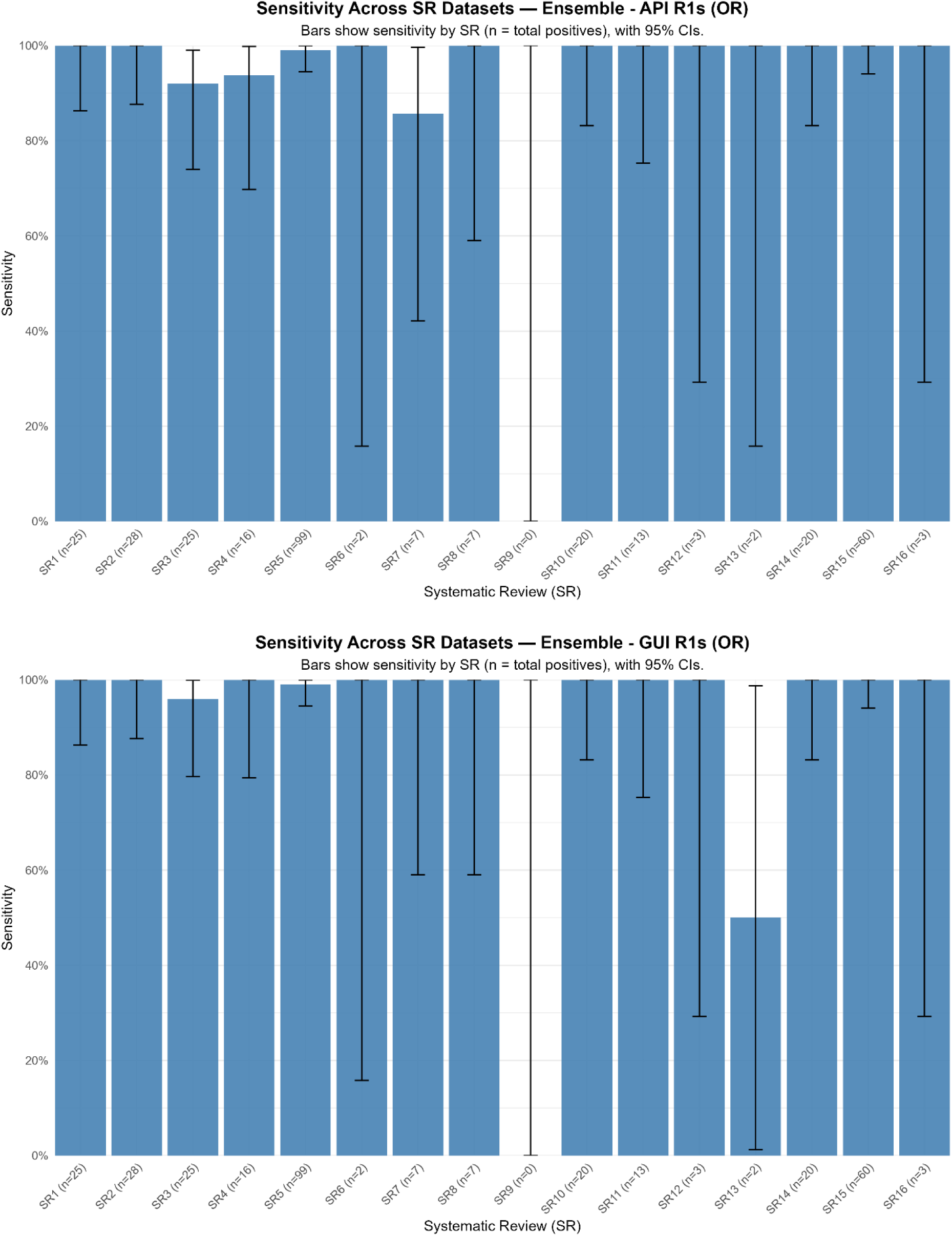
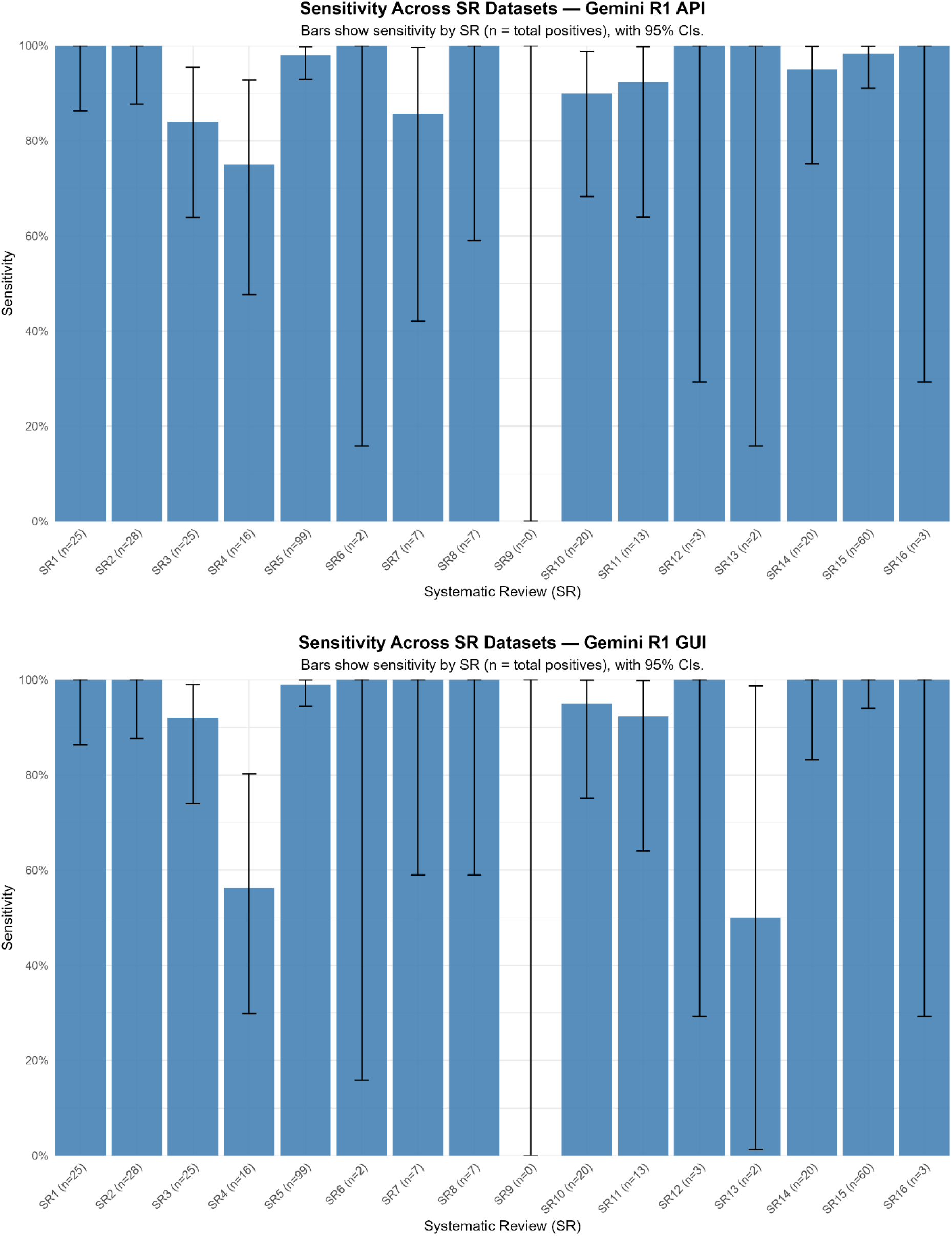
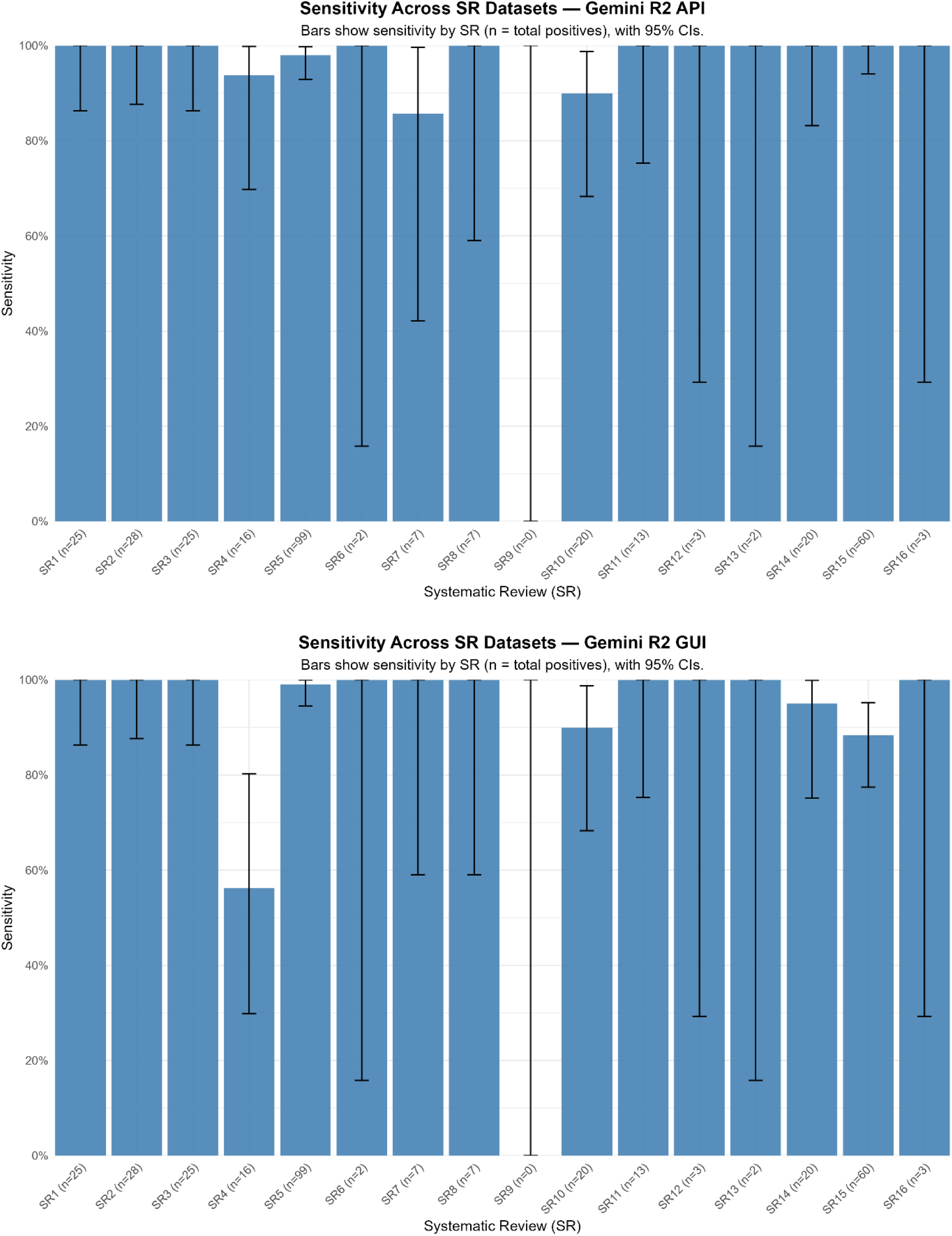
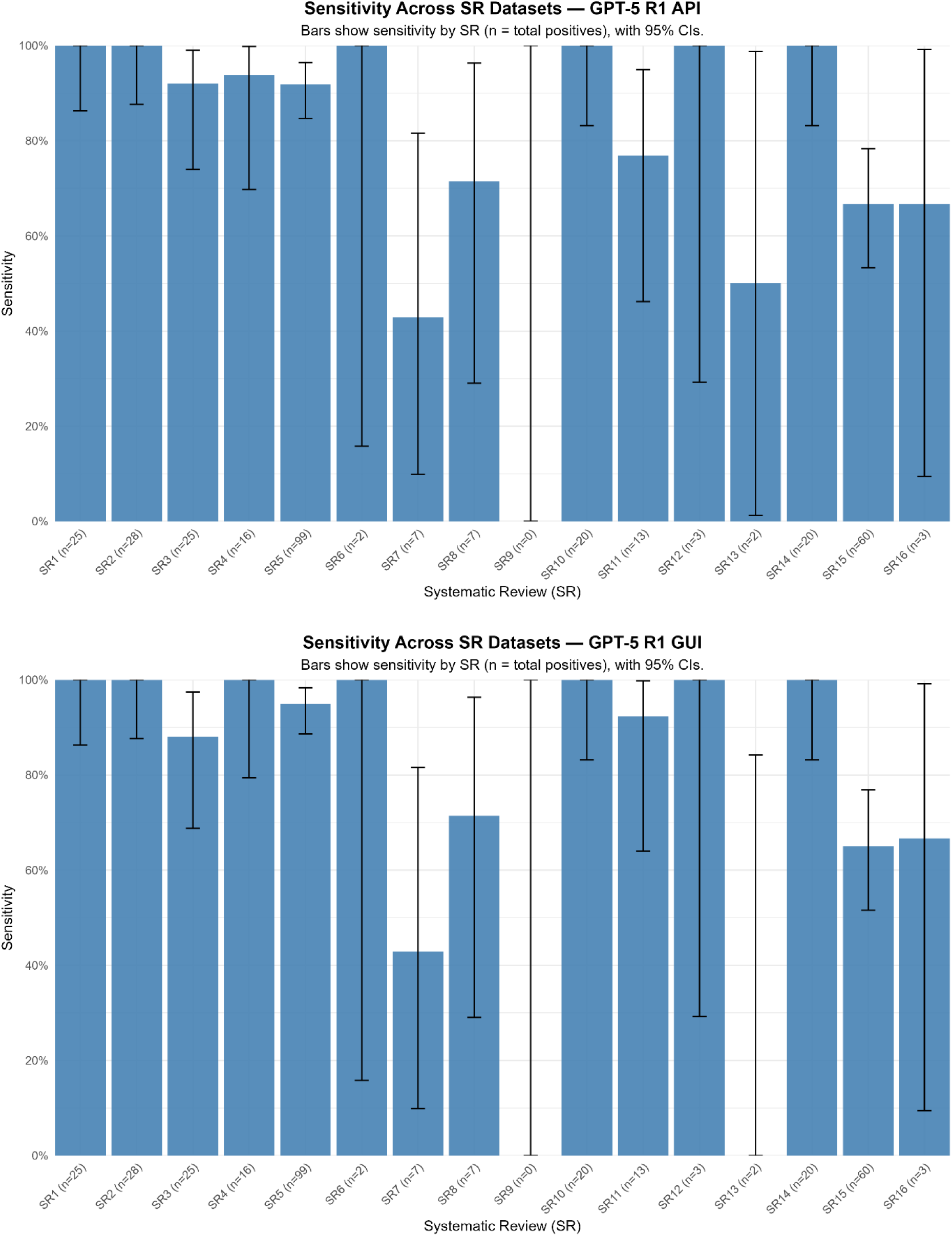
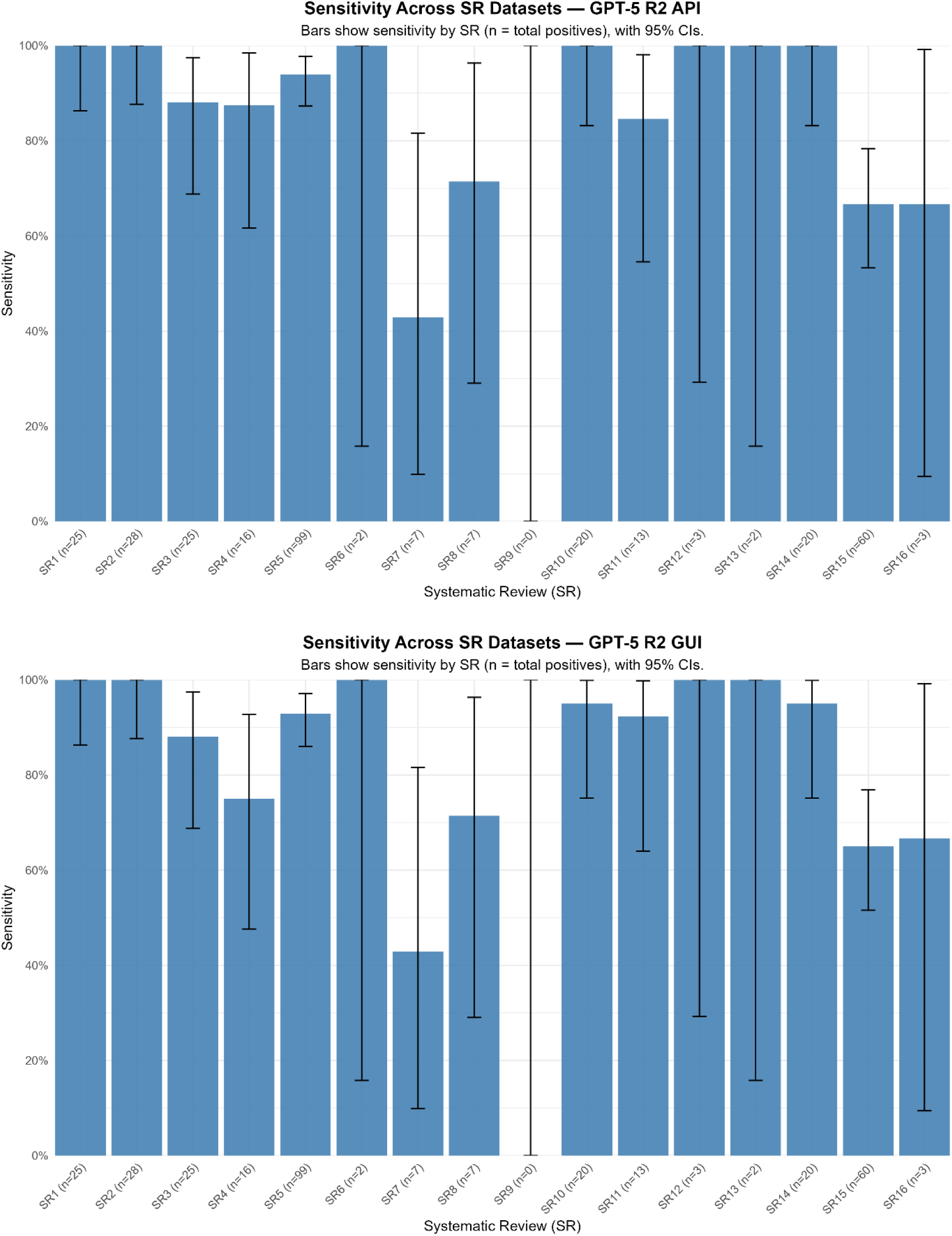
Per-Review Sensitivity for Each Variant. Bar charts showing the sensitivity for each of the 16 systematic reviews (SRs) for a specific model variant. Error bars represent 95% Clopper-Pearson confidence intervals. The number of positive records (n) for each SR is shown on the x-axis.

## LLM Performance vs. Original Cochrane EndNote library

**Table S2.** Overall Performance of All Model Variants. Summary of performance metrics, including true positives (TP), false negatives (FN), true negatives (TN), false positives (FP), sensitivity, and specificity. Confidence intervals (CI) are 95% Clopper-Pearson intervals.

| LLM | TP | FN | TN | FP | Sensitivity (95% CI) | Specificity (95% CI) |
| --- | --- | --- | --- | --- | --- | --- |
| **GPT-5 R1 API** | 288 | 60 | 246 | 142 | 82.8% (78.4%, 86.6%) | 63.4% (58.4%, 68.2%) |
| **GPT-5 R2 API** | 290 | 58 | 246 | 142 | 83.3% (79.0%, 87.1%) | 63.4% (58.4%, 68.2%) |
| **GPT-5 R1 GUI** | 291 | 57 | 254 | 134 | 83.6% (79.3%, 87.4%) | 65.5% (60.5%, 70.2%) |
| **GPT-5 R2 GUI** | 285 | 63 | 246 | 142 | 81.9% (77.4%, 85.8%) | 63.4% (58.4%, 68.2%) |
| **Gemini R1 API** | 314 | 34 | 222 | 166 | 90.2% (86.6%, 93.1%) | 57.2% (52.1%, 62.2%) |
| **Gemini R2 API** | 324 | 24 | 222 | 166 | 93.1% (89.9%, 95.5%) | 57.2% (52.1%, 62.2%) |
| **Gemini R1 GUI** | 317 | 31 | 235 | 153 | 91.1% (87.6%, 93.9%) | 60.6% (55.5%, 65.5%) |
| **Gemini R2 GUI** | 312 | 36 | 238 | 150 | 89.7% (86.0%, 92.6%) | 61.3% (56.3%, 66.2%) |
| **Ensemble - Any GUI (OR)** | 329 | 19 | 182 | 206 | 94.5% (91.6%, 96.7%) | 46.9% (41.9%, 52.0%) |
| **Ensemble - Any API (OR)** | 327 | 21 | 182 | 206 | 94.0% (90.9%, 96.2%) | 46.9% (41.9%, 52.0%) |
| **Ensemble - GUI R1s (OR)** | 327 | 21 | 210 | 178 | 94.0% (90.9%, 96.2%) | 54.1% (49.0%, 59.2%) |
| **Ensemble - API R1s (OR)** | 325 | 23 | 201 | 187 | 93.4% (90.2%, 95.8%) | 51.8% (46.7%, 56.9%) |

**Supplementary Figure S13-S24:**
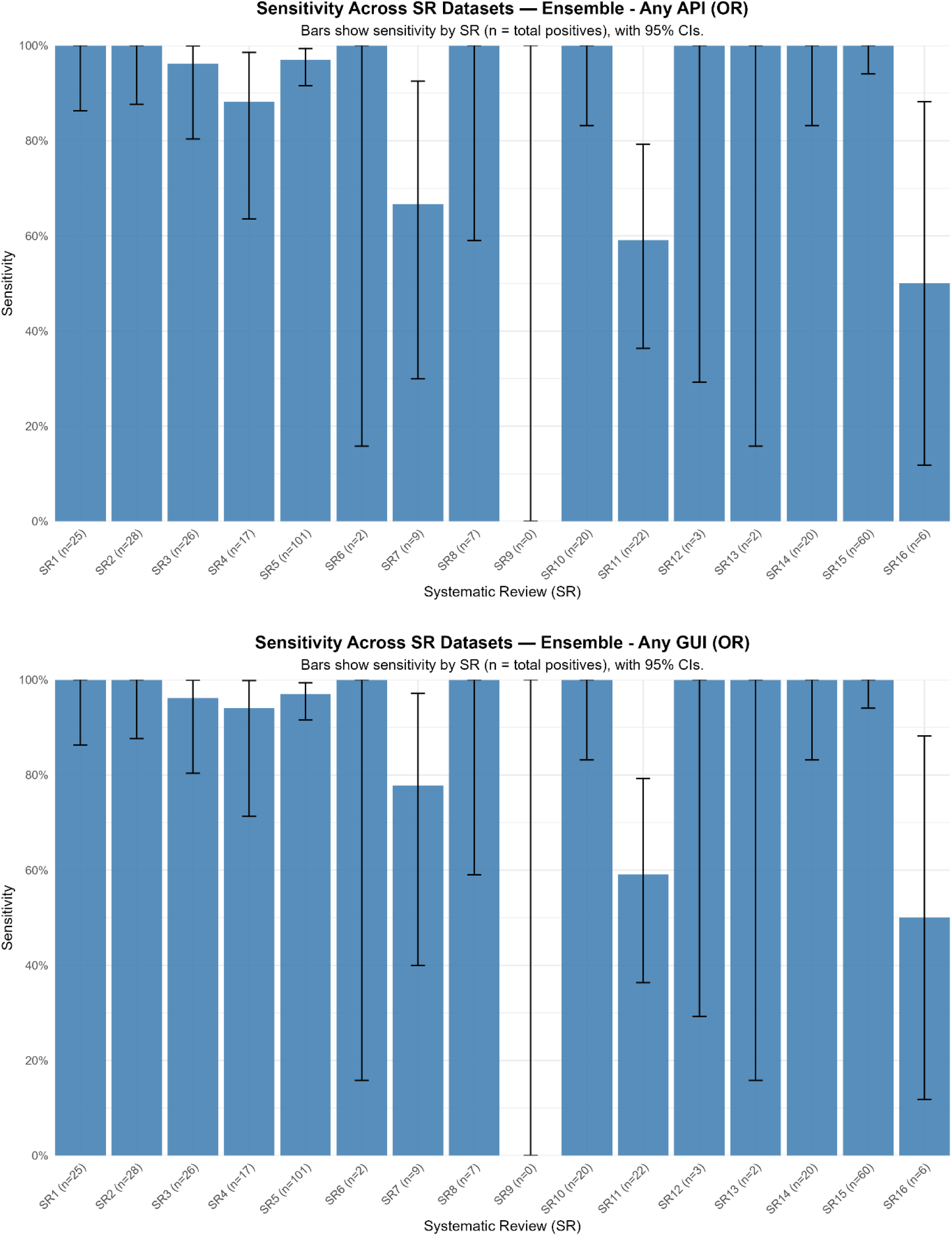
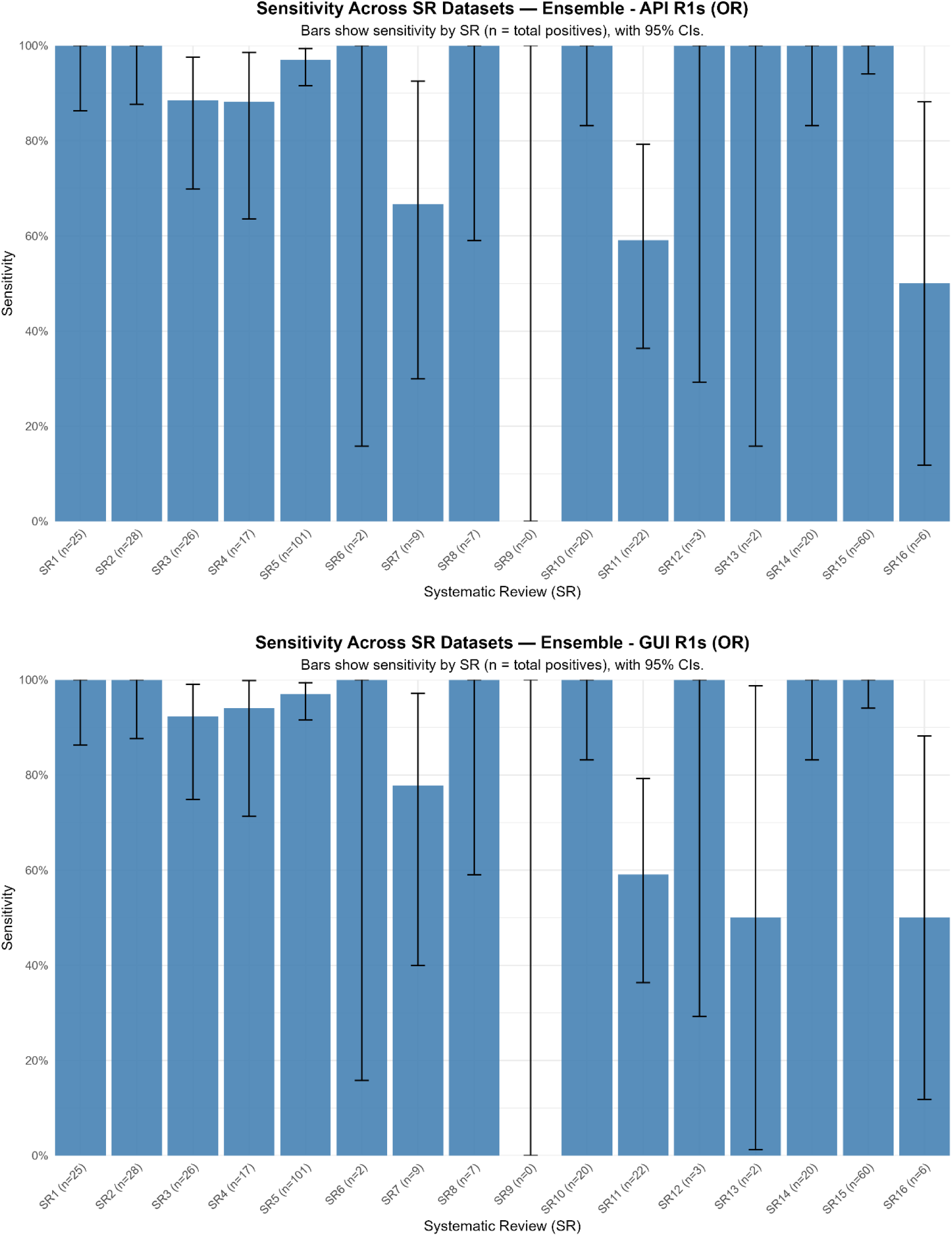
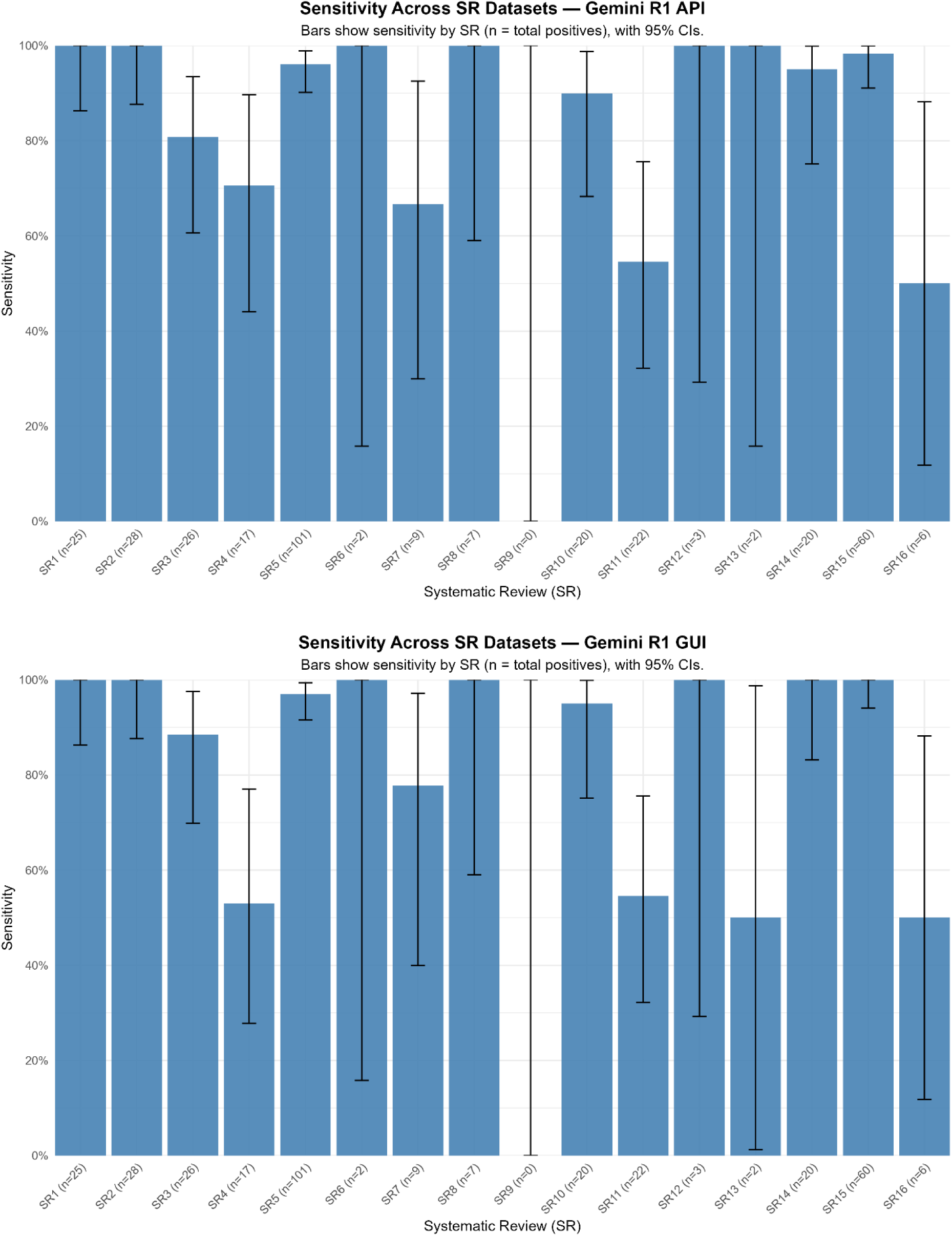
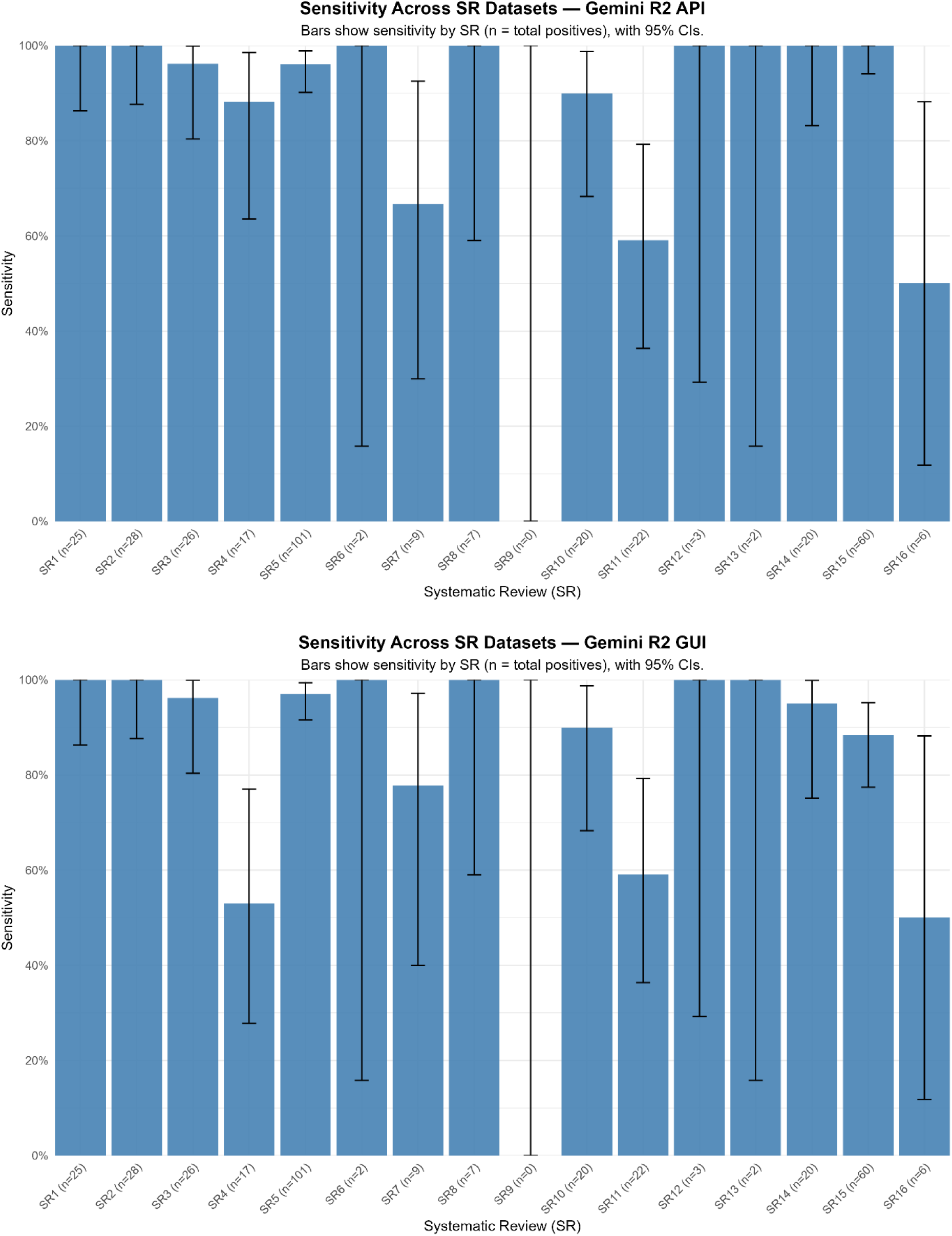
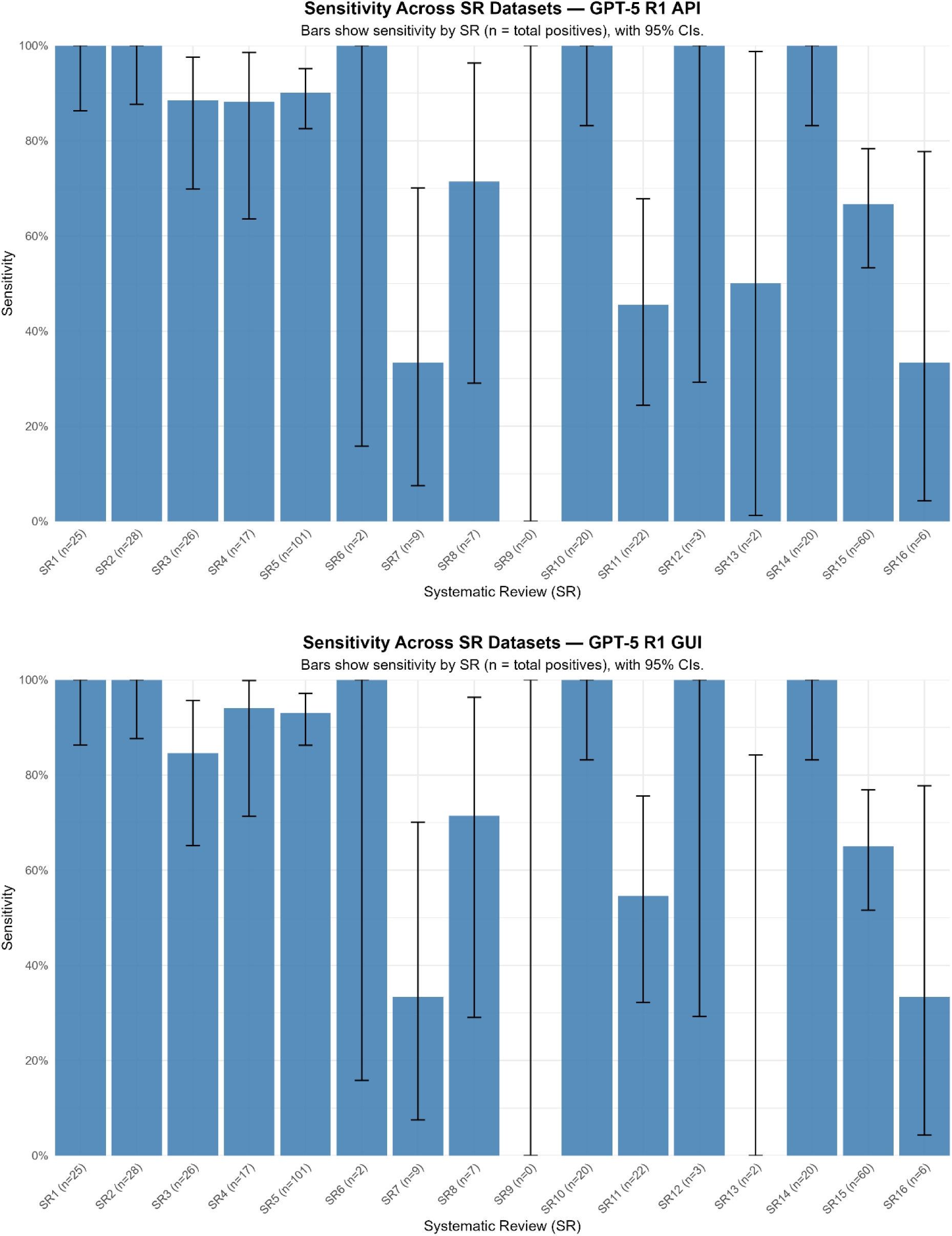
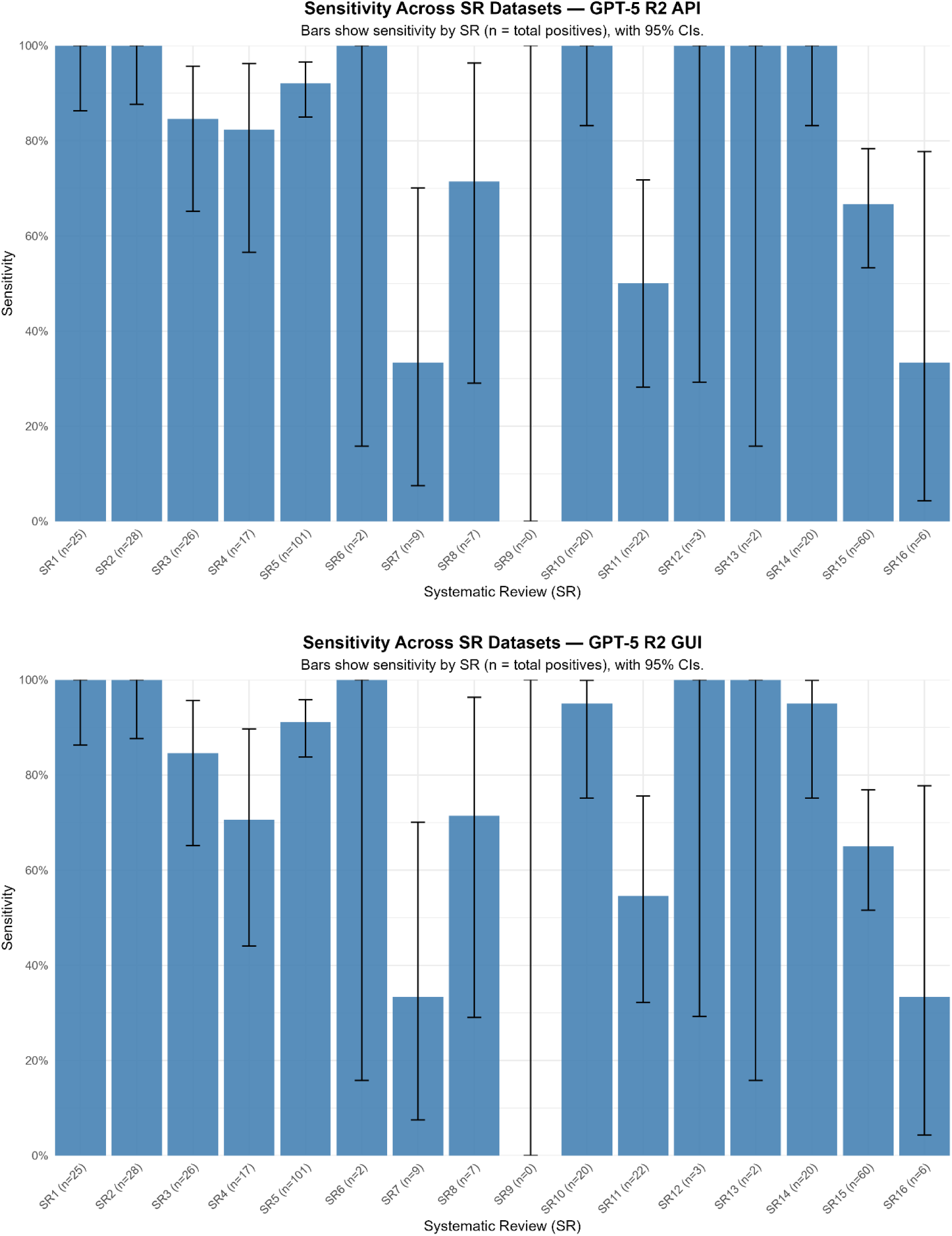
Per-Review Sensitivity for Each Variant. Bar charts showing the sensitivity for each of the 16 systematic reviews (SRs) for a specific model variant. Error bars represent 95% Clopper-Pearson confidence intervals. The number of positive records (n) for each SR is shown on the x-axis.

